# Direct and indirect effects of the COVID-19 pandemic on mortality in Switzerland: A population-based study

**DOI:** 10.1101/2022.08.05.22278458

**Authors:** Julien Riou, Anthony Hauser, Anna Fesser, Christian L. Althaus, Matthias Egger, Garyfallos Konstantinoudis

**Affiliations:** Institute of Social and Preventive Medicine, University of Bern, Switzerland; Federal Office of Public Health, Switzerland; Population Health Sciences, Bristol Medical School, University of Bristol, Bristol, UK; Centre for Infectious Disease Epidemiology and Research, University of Cape Town, Cape Town, South Africa; MRC Centre for Environment and Health, Department of Epidemiology and Biostatistics, School of Public Health, Imperial College London, London, UK

## Abstract

The direct and indirect impact of the COVID-19 pandemic on population-level mortality is of concern to public health but challenging to quantify. We modelled excess mortality and the direct and indirect effects of the pandemic on mortality in Switzerland. We analyzed yearly population data and weekly all-cause deaths by age, sex, and canton 2010-2019 and all-cause and laboratory-confirmed COVID-19 deaths from February 2020 to April 2022 (study period). Bayesian models predicted the expected number of deaths. A total of 13,130 laboratory-confirmed COVID-19 deaths were reported. The model estimated that COVID-19-related mortality was underestimated by a factor of 0.72 [95% Credible Interval: 0.46-0.78] resulting in 18,140 [15,962-20,174] excess deaths. After accounting for COVID-19 deaths, the observed mortality was 3% [-1-7] lower than expected, corresponding to a deficit of 4,406 deaths, with a wide credibility interval [-1,776-10,700]. Underestimation of COVID-19 deaths was greatest for ages 70 years and older; the mortality deficit was most pronounced in age groups 40 to 69 years. We conclude that shortcomings in testing caused underestimation of COVID-19-related deaths in Switzerland, particularly in older people. Although COVID-19 control measures may have negative effects (e.g., delays in seeking care or mental health impairments), after subtracting COVID-19 deaths, there were fewer deaths in Switzerland during the pandemic than expected, suggesting that any negative effects of control measures on mortality were offset by the positive effects. These results have important implications for the ongoing debate about the appropriateness of COVID-19 control measures.

## Introduction

The COVID-19 pandemic has affected mortality globally through direct and indirect effects. Infection with SARS-CoV-2 directly causes death in a small proportion of infected people, with variations in the infection-fatality ratio (IFR) depending on age (1), socio-economic position (2), vaccination status (3), intensive care unit capacity (4) and several other characteristics of individuals or communities. Combined with the high transmissibility of SARS-CoV-2, this resulted in more than 6 million laboratory-confirmed deaths globally as of July 12, 2022 (5). The pandemic has also caused major disruptions in many aspects of social and economic life and may thus have indirectly increased or reduced mortality. Non-pharmaceutical interventions (NPIs) may lead to delays or avoidance of medical care (6, 7), increases in substance use and suicidal ideation (8-10), or increases in interpersonal violence (11). Conversely, stay-at-home orders have led to reductions in mobility and traffic (12) and air pollution levels (13). Border closures and cutbacks in social contacts and activities have restricted the circulation of other infectious diseases (14). The respective importance of the pandemic’s positive and negative indirect effects on mortality and the net impact remains unknown.

The overall impact of the COVID-19 pandemic on mortality at the population level, both directly and indirectly, is of great concern to public health but is difficult to quantify. Laboratory-confirmed deaths (i.e., deceased people with a recent positive SARS-CoV-2 test) may underestimate mortality as some deaths will remain unascertained, for example, due to testing policies, shortages, underreporting, or overwhelmed health systems (15). Laboratory-confirmed deaths ignore indirect effects on mortality. The main alternative relies on excess mortality estimated from all-cause mortality data, using counterfactual reasoning (16). The observed number of deaths is compared to what would have been expected had the pandemic not occurred, based on mortality data from previous years and considering demographic changes and covariates associated with mortality patterns. The approach has the advantage of covering both the pandemic’s direct and indirect effects, although phenomena like mortality displacement can limit the interpretability of results (17-19). Also, estimations of excess mortality depend on model assumptions and methodological choices, such as age-specific population trends (20). Detailed analyses of causes of death as listed in death certificates can also be used (generally with considerable delay) but suffer from significant limitations, especially regarding ascertaining infectious diseases (21).There have been many attempts to estimate excess mortality associated with the COVID-19 pandemic (22-32). Comparisons of excess mortality with laboratory-confirmed deaths have confirmed that the overall impact of the pandemic on mortality is generally much greater than what is indicated by laboratory-confirmed deaths alone (23, 28, 30). Still, a common limitation of these studies is the inability to distinguish between the direct and indirect effects of the pandemic on mortality. This study attempts to overcome this limitation by jointly studying laboratory-confirmed COVID-19-related deaths and excess mortality. We computed the expected number of all-cause deaths by week, age group, and location in Switzerland between February 2020 and April 2022, accounting for the effect of temperature, national holidays, and population changes using a validated statistical approach (33). We then developed a method to decompose all-cause mortality into deaths directly attributable to SARS-CoV-2 infection and deaths indirectly attributable to the pandemic. We could thus examine the completeness of ascertainment of COVID-19-related deaths and the indirect effects of the pandemic on all-cause mortality in Switzerland.

## Methods

### Data sources

We retrieved population data in Switzerland for the pre-pandemic years 2010 to 2019 from the *Federal Statistical Office* (FSO). Data were aggregated by age group (in five groups: 0-39, 40-59, 60-69, 70-79 and 80 and older), sex (two groups) and administrative region (26 cantons). Data on all-cause deaths were also obtained from the FSO. These consisted of counts of deaths from any cause by age, sex, and canton for each week from 2010 to 2019, and afterwards for each week up to April 3, 2022. Coding the cause of death listed in the death certificate is time-consuming. Therefore, data on cause of death are published up to 24 months after a completed calendar year and were therefore not available for this analysis. We used data on ambient temperature from the European Centre for Medium-Range Weather Forecasts Reanalysis version 5 (ERA5) reanalysis data set (34) and on national holidays. Daily mean ambient temperature between 2010 and 2022 at 0.25°x0.25° resolution was aggregated by taking means per week and canton. Holidays were defined weekly for each canton (1 if there was at least one cantonal holiday, 0 otherwise). The reporting of laboratory-confirmed COVID-19-related deaths has been mandatory in Switzerland since February 2020. The records are kept at the *Federal Office of Public Health* (FOPH) and are available online. Data include age, sex, canton of residence, and the date and type of the positive SARS-CoV-2 test. Dates were grouped into seven epidemic phases by the FOPH: February 24, 2020 to June 7, 2020 (phase 1); June 8, 2020 to September 27, 2020 (phase 2); September 28, 2020 to February 14, 2021 (phase 3); February 15, 2021 to June 20, 2021 (phase 4); June 21, 2021 to October 10, 2021 (phase 5); October 11, 2021 to December 19, 2021 (phase 6) and December 20, 2021 to April 3, 2022 (phase 7).

### Population trends model

We used population size on December 31, 2010 to 2019 by age group, sex and canton to predict population sizes in each stratum and week of the entire study period (January 1, 2020 to April 3, 2022) in a two-step procedure. First, we fitted a Poisson regression model to population data from 2010 to 2019. This model included a linear yearly trend, a fixed effect by sex, and independent random effects by week (for seasonality), age group and canton. We compared different models using higher interactions and yearly linear trends that vary by space, age, and sex. Model comparison using a cross-validation scheme excluding the last three years of available data (2017-2019) determined that the best model included all possible two-way interactions between age, canton, and week, and an overdispersion parameter. We obtained posterior distributions of the population in each stratum for December 31 2020, 2021 and 2022, under the counterfactual scenario that the pandemic did not occur. In a second step, we used linear interpolation to obtain weekly population size (estimates, with uncertainty). SI Appendix Section A provides further details.

### Expected deaths model

We estimated the expected number of all-cause deaths for each week between February 24, 2020, the day of the first confirmed COVID-19 case in Switzerland, and April 3, 2022 by age, sex and canton of residence using the historical data (2011 to 2019) and expanding a previously proposed model (25). We used Bayesian spatio-temporal models accounting for population trends and including covariates related to temperature and national holidays. To account for uncertainty in population estimates, we applied the model multiple times over the samples of the posterior distributions of the population predictions. Since the effect of temperature on all-cause mortality is expected to be U-shaped (35), we used a random walk of order 2 to allow for a flexible fit. We accounted for seasonality using a random walk of order 1 at the weekly level, and for exceptional events using week-level independent random effects. We accounted for long-term trends with a linear slope at the yearly level, and for spatial autocorrelation using conditional autoregressive priors. We modelled spatial autocorrelation using an extension of the Besag-York-Mollié model, allowing for a mixing parameter measuring the proportion of the marginal variance explained by the spatial autocorrelation term (36, 37). The model has been internally validated and found to be unbiased and to have a high predictive accuracy in age groups above 40. We used the fitted model to obtain posterior distributions of the expected number of all-cause deaths by age group, sex and canton in each week between February 24, 2020 and April 3, 2022. Estimates of excess mortality (with uncertainty) were then obtained by subtracting the expected (across the posterior samples) from the observed all-cause deaths in each stratum. SI Appendix Section B provides further details and the results of the internal cross validation.

### Decomposition model

We first studied the alignment between excess mortality and laboratory-confirmed COVID-19-related deaths using Pearson’s correlation coefficient (applied across the posterior samples of excess mortality to propagate uncertainty). We then developed a method to decompose the number of all-cause deaths observed in the pandemic period based on 1) the number of laboratory-confirmed COVID-19-related deaths and 2) the number of expected deaths given historical trends. We included multiplicative parameters to measure the respective contributions of these two quantities. We used a Poisson regression model with an identity link and no intercept term of the form:

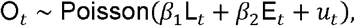

where *O*_*t*_ is the observed number of all-cause deaths on week *t, L*_*t*_ is the number of laboratory-confirmed COVID-19-related deaths, *E*_*t*_ is the expected number of all-cause deaths given historical trends, and *u*_*t*_ is a normally distributed overdispersion term centered at zero. Within this formulation, *β*_l_ is the number of all-cause deaths for each unit increase in laboratory-confirmed deaths, after adjusting for the expected number of all-cause deaths given historical trends. That means that under perfect case ascertainment *β*_l_ *= 1*. If *β*_l_ > 1, then we observe a greater number of deaths attributed to SARS-CoV-2 infections compared with the number of laboratory-confirmed deaths. The ascertainment proportion of COVID-19-related deaths is obtained by 1/*β*_l_. This relies on the assumption that when there is at least one laboratory-confirmed death in a week, then the excess in observed all-cause deaths can be directly attributed to COVID-19. In a similar way, *β*_2_ is the number of all-cause deaths for each unit increase in the expected number of all-cause deaths after adjusting for the direct effect of COVID-19. We expect *β*_2_ =1 when the net effect of the pandemic-related behavioral, societal and health system changes on all-cause deaths is zero. The estimate of *β*_2_ can thus be interpreted as a measure of the indirect effect of the pandemic on mortality. If *β*_2_ < 1, then there were fewer all-cause deaths than expected after removing the direct effect of COVID-19, which could imply an indirect protective effect of all changes and control measures associated with the pandemic. Estimates of *β*_l_ and *β*_2_ thus provide a way to understand the interplay between laboratory-confirmed COVID-19-related deaths and excess all-cause deaths and allow to differentiate between direct and indirect consequences of the COVID-19 pandemic on mortality. SI Appendix Section C provides further details on model specification and choices of the priors.

We extended the model presented above to examine these associations by phase (from 1 to 7 as defined by the FOPH), by age group (0-39, 40-59, 60-69, 70-79 and 80+ years old), and by area (26 cantons). To this end, we introduced multiple *β*_l_ and *β*_2_ for each phase, age group or area separately, with the additional constraint of a multilevel structure allowing a smoothing towards the global mean of the estimator (38). To propagate the uncertainty of the expected number of deaths, we fitted the above models using 200 samples of the posterior distribution of the expected number of deaths. We then combined the resulting posterior samples of *β*_l_ and *β*_2_. All inferences were done in a Bayesian framework. Posterior distributions were approximated by samples, and summarized by their median, 2.5% and 97.5% percentiles to obtain point estimates and 95% credible intervals (95% CrI). The population and expected deaths models were implemented in R-INLA (39), and the decomposition model in NIMBLE (40). The code is available on github at https://github.com/jriou/covid19_ascertain_deaths.

## Results

We observed a total of 156,193 deaths from all causes in Switzerland from February 24, 2020, to April 3, 2022, compared to an expected 142,408 [95% Credible Interval: 138,044-149,125] had the pandemic not occurred. This translates into 13,785 [7,068-18,149] excess all-cause deaths over the pandemic period, a relative increase of 9.7% [4.7-13.1]. There were three periods of substantial relative excess mortality: 7.3% [3.8-10.8] during phase 1, 33.9% [26.4-41.4] during phase 3 and 15.9% [8.3-22.8] during phase 6 (Table 1). There was some evidence suggesting mortality displacement during phase 4, with a relative excess mortality of -4.3% [-9.9-0.2]. The age groups affected most by excess mortality were those over 70 years of age (Figure 1A and B). A total of 13,130 laboratory-confirmed COVID-19-related deaths were reported during the study period. Weekly counts of laboratory-confirmed deaths generally aligned with estimates of excess all-cause mortality in Switzerland (Figure 2), with a correlation coefficient of 0.89 [0.85-0.92]. The estimate of *β*_l_ was 1.38 [1.22-1.54], suggesting that there were, on average, 38% [22-54] more deaths directly attributable to COVID-19 than laboratory-confirmed deaths during the period or that the ascertainment proportion was 72% [65-82] (Table 1). Given the 13,130 laboratory-confirmed deaths over the period, this implies that the total number of deaths directly attributable to COVID-19 in Switzerland until April 3, 2022, can be estimated at 18,140 [15,962-20,174] deaths.

**Table 1.** Mean and 95% credible intervals for expected and excess number of all-cause deaths, relative excess all-cause mortality, and number of observed all-cause and laboratory-confirmed COVID-19-related deaths by seven epidemic phases between February 2020 to April 2022.

| Phase* | Expected | Observed | Excess | Relative excess | Laboratory |
| --- | --- | --- | --- | --- | --- |
| 1 | 19,376<br>(18,767 to 20,033) | 20,791 | 1,415<br>(758 to 2,024) | 7%<br>(4 to 11) | 1,725 |
| 2 | 19,180<br>(18,440 to 20,042) | 19,103 | -76<br>(-939 to 663) | -0%<br>(-5 to 4) | 104 |
| 3 | 27,004<br>(25,569 to 28,604) | 36,157 | 9,154<br>(7,553 to 10,588) | 34%<br>(26 to 41) | 7,652 |
| 4 | 23,386<br>(22,320 to 24,834) | 22,369 | -1,017<br>(-2,465 to 49) | -4%<br>(-10 to 0) | 895 |
| 5 | 19,174<br>(18,284 to 20,223) | 20,007 | 832<br>(-216 to 1,723) | 4%<br>(-1 to 9) | 380 |
| 6 | 13,036<br>(12,298 to 13,944) | 15,105 | 2,070<br>(1,161 to 2,807) | 16%<br>(8 to 23) | 956 |
| 7 | 21,370<br>(20,067 to 22,894) | 22,661 | 1,291<br>(-233 to 2,594) | 6%<br>(-1 to 13) | 1,418 |

\* Phase 1: February 24, 2020 to June 7, 2020, phase 2: June 8, 2020 to September 27, 2020, phase 3: September 28, 2020 to February 14, 2021, phase 4: February 15, 2021 to June 20, 2021, phase 5: June 21, 2021 to October 10, 2021, phase 6: October 11, 2021 to December 19, 2021 and phase 7: December 20, 2021 to April 3, 2022.

**Figure 1.**
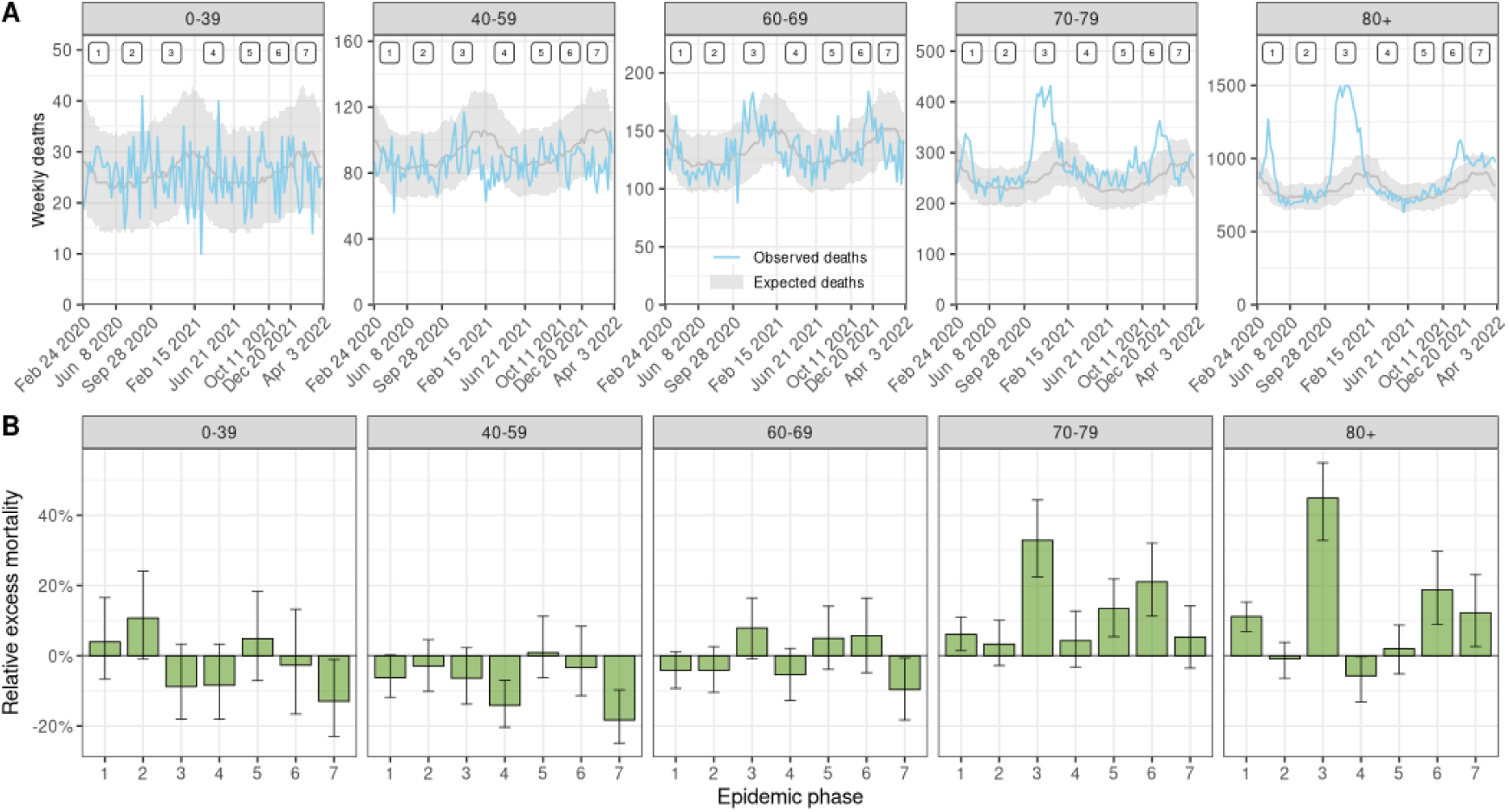
(A) Observed and expected number of weekly deaths by age group in Switzerland from February 2020 to April 2022. Model-predicted expected deaths are shown with median and 95% credibility interval. Numbers at the top indicate epidemic phases 1 to 7. (B) Estimated relative excess mortality by seven epidemic phases from February 2020 to April 2022 and five age groups. Medians with 95% credible intervals are shown.

**Figure 2.**
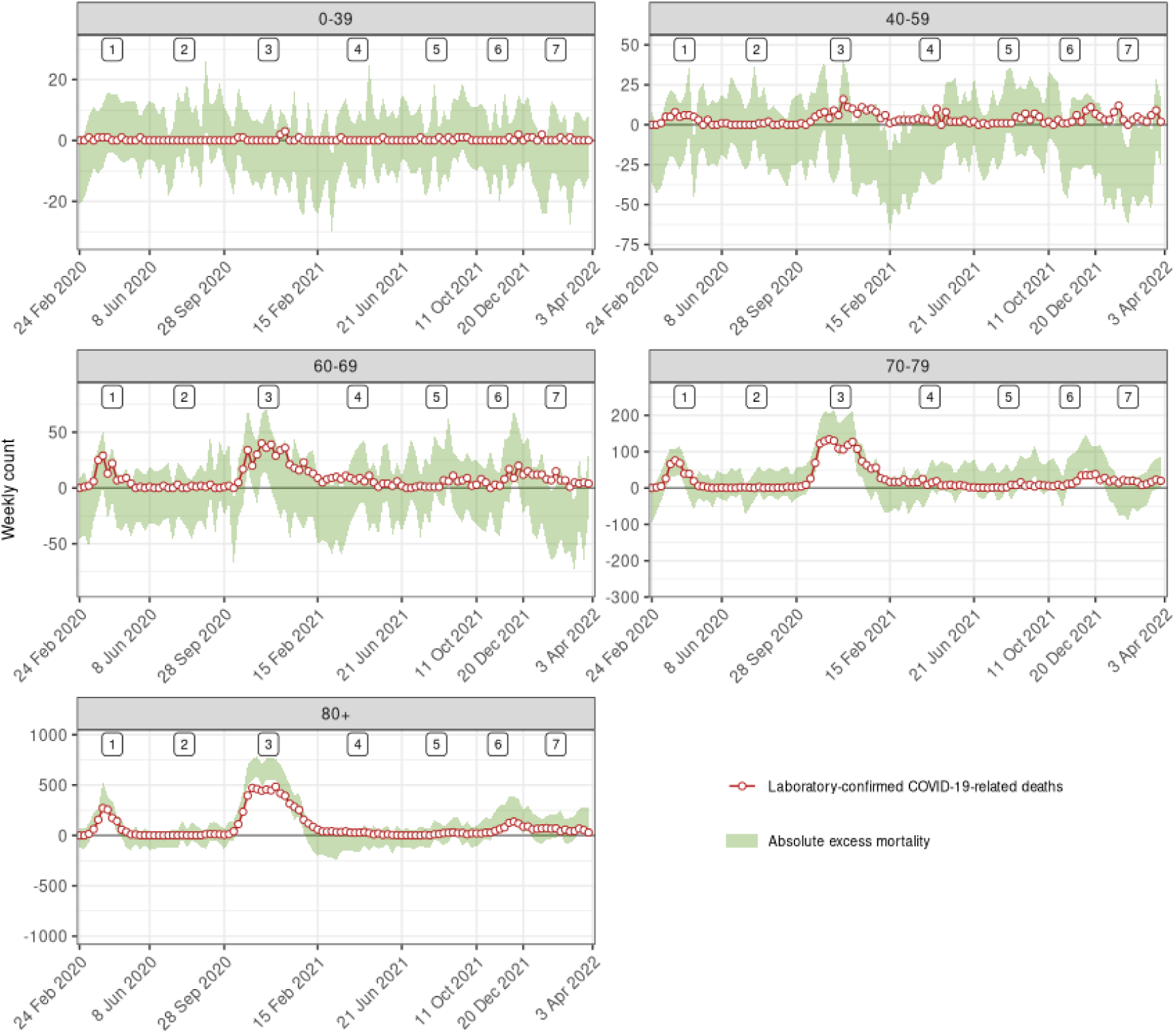
Weekly counts of excess all-cause deaths (95% credibility intervals) and of laboratory-confirmed COVID-19-related deaths between February 24, 2020 and April 3, 2022 in Switzerland by five age groups. Numbers at the top indicate epidemic phases 1 to 7.

After accounting for deaths directly attributable to COVID-19, the observed number of all-cause deaths was slightly lower than expected based on historical trends. This deficit is quantified by *β*_2_, estimated at 0.97 [0.93-1.01], indicating 3% [-1-7] fewer all-cause deaths than expected during the COVID-19 pandemic after adjusting for the direct effect of SARS-CoV-2 infections on mortality. This corresponds to 4,406 [-1,776-10,700] fewer deaths overall compared to expected. The credibility interval is wide, indicating that the data are compatible with no indirect beneficial effect or a slightly harmful indirect effect.

The coefficients *β*_l_ and *β*_2_ varied across age groups and time periods. The alignment between excess mortality and laboratory-confirmed deaths was particularly noticeable in age groups 70-79 and 80 and older (Figure 3A and SI Appendix Table S1), and during phases 1, 3 and 6 (SI Appendix, Figure S1). Variation in the relative number of deaths directly attributable to COVID-19 for each laboratory-confirmed death (*β*_l_) by age group suggests that more deaths were not ascertained in age groups 70-79 and 80+, while the data were compatible with 100% ascertainment (*β*_l_ = 1) in age groups below 70, where fewer deaths were reported (Figure 3B). *β*_l_ was estimated around 1.5 during phases 1 and 3 and around 2 during phase 6, suggesting an ascertainment proportion of COVID-19 deaths during large epidemic waves ranging between 50 and 66% (Figure 3B). This estimate is less precise during periods of low epidemic activity (phases 2, 4, 5 and 7), and remains compatible with 1 (perfect ascertainment). The relative deficit in all-cause deaths (*β*_2_) was more pronounced in age groups 40 to 69 and during phases 1, 3 and 4 (Figure 3C). Estimates of *β*_l_ and *β*_2_ across administrative regions show generally homogeneous results for the whole of Switzerland (SI Appendix, Figure S2 and Table S2).

**Figure 3.**
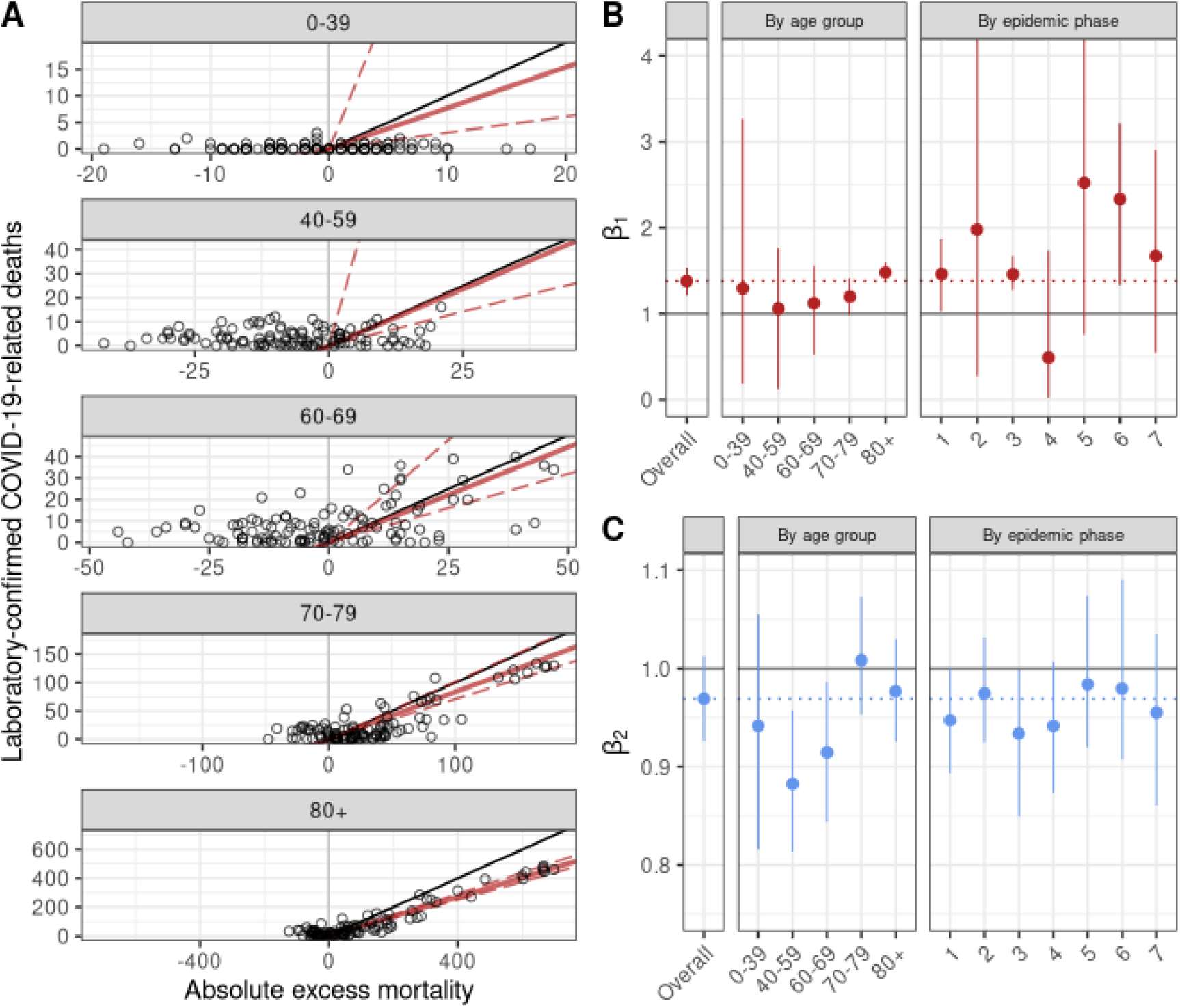
(A) Association between weekly laboratory-confirmed COVID-19-related deaths and absolute excess mortality by age group. The black line shows the slope of association corresponding to a 1 to 1 relation. The red lines show the association estimated with the model (corresponding to the *β*_l_ coefficients shown in panel B, the full line represents the point estimate and the dashed lines the lower and upper bounds of the 95% credible interval). (B) Estimates of *β*_l_, the additional number of deaths to be observed for each unit increase in laboratory-confirmed deaths, after adjusting for the expected number of all-causes deaths given historical trends. (C) Estimates of *β*_2_, the additional number of deaths to be observed for each unit increase in the expected number of all-cause deaths, after adjusting for the direct effect of SARS-CoV-2 infections. Estimates of *β*_l_ and *β*_2_ are shown for the whole period, by phase and by age group.

## Discussion

We examined the patterns of all-cause mortality in Switzerland from the diagnosis of the first COVID-19 case at the end of February 2020 to spring 2022. We compared the excess mortality with laboratory-confirmed COVID-19-related deaths and separated excess mortality into the excess directly attributable to COVID-19 and excess or under mortality indirectly attributable to the pandemic. We found that the estimated number of deaths directly caused by COVID-19 was about 1.4 times higher than the number of laboratory-confirmed deaths. In other words, only about 70% of COVID-19-related deaths were ascertained. Overall, COVID-19 was directly responsible for an estimated 18,000 deaths during the study period, during which only around 13,000 laboratory-confirmed COVID-19-related deaths were reported. Finally, the pandemic may have had an indirect beneficial effect on mortality, estimated at around 4,000 fewer deaths than expected. Of note, this protective effect primarily concerned younger age groups.

Our study has several strengths. Detailed data on the population structure, mortality, weather, and national holidays from the ten years before the COVID-19 pandemic allowed us to estimate what mortality would have been 2020 to 2022 had the pandemic not occurred. Estimation of excess all-cause mortality during the pandemic period by time, space and age thus became possible. Our method has been thoroughly validated (SI Appendix Supporting Information Text) and accounts for the most important determinants of all-cause mortality, including projected population sizes and observed temperature. It correctly handles uncertainty from the different data sources and propagates it to the final estimates. The approach may be used in any setting with reliable reports of all-cause mortality and laboratory-confirmed deaths. Further, we developed a novel statistical method to differentiate between deaths directly attributable to SARS-CoV-2 infections and deaths caused or prevented indirectly by the pandemic. Our study also has several limitations. Information about the cause of death during the study period was not available but would have helped understand the mechanisms of the indirect beneficial effect of the COVID-19 pandemic on mortality. We assumed deaths with a positive SARS-CoV-2 test were caused by COVID-19, although the infection could be incidental in some cases. However, incidental SARS-CoV-2 infections would only concern a small proportion of deaths with laboratory-confirmed infection, equivalent to the prevalence in the general population (41). Autopsies of patients dying in hospital following a positive SARS-CoV-2 test also suggest that during the study period, causes of death were generally directly related to COVID-19 (42). Finally, we did not stratify by sex in this study. In a previous analysis, we found only small differences in excess deaths between sexes (25).

Our estimates of overall excess mortality during the COVID-19 pandemic in Switzerland are consistent with other analyses. The Federal Office of Statistics reported an excess mortality above 10% for January 2020 to August 2021 (29), higher than the 9.7% estimated in the present study for the period up to spring 2022. A multi-country study estimated excess mortality at 13,000 deaths for Switzerland during March 2020 to June 2022 (24, 43) and another one at 15,500 [14,000-17,000] for 2020 and 2021 (28). In our study of five European countries we estimated an excess mortality of 8% in males and 9% in females for Switzerland during the first year of the pandemic (25). WHO estimates for Switzerland 2020 and 2021 were lower (8,200 excess deaths), but there were problems with the WHO estimates (26, 44). None of these studies attempted to quantify the direct and indirect effects of the pandemic on mortality. In line with our estimates, a study in California reported a 78% ascertainment proportion of diagnosed COVID-19 deaths, but did not quantify the indirect effect of the pandemic (45). This is the first study that aims to quantify both the direct and indirect effects of the pandemic on mortality.

We found that COVID-19 caused about 1.4 times more deaths than were laboratory-confirmed, in line with a recent study estimating this ratio at 1.29 [1.16-1.42] for Switzerland (30). Estimates varied widely between countries. For example, the ratio was 0.57 [0-1.25] for Norway but at 150 [140-162] for Nicaragua (30). Differences could be attributable to local healthcare and surveillance systems, testing capacity, and methodological differences in collecting mortality data and estimating excess mortality. We found markedly lower ascertainment during periods of high epidemic activity, suggesting shortcomings in testing, or reporting even in Switzerland, a high-income country. Under-ascertainment was concentrated in older age groups, indicating incomplete ascertainment in retirement and nursing homes, in line with other reports (15). The lower ascertainment towards the end of the study period might be explained by reduced testing once vaccines became available.

We found that in Switzerland, the COVID-19 pandemic probably had an indirect beneficial effect on mortality. A potential explanation is mortality displacement or the “harvesting effect”, where COVID-19 precipitated deaths that would have occurred anyway soon (17-19, 46). However, this is unlikely: the deficit of deaths was concentrated in the younger age groups and not in the over 70 years old, where mortality displacement typically occurs. Therefore, the mortality deficit is probably due to the pandemic’s indirect effects, such as reductions in mobility, road traffic, air pollution and sports activities. The fact that the deficit was more pronounced during phases 1, 3 and 4, when control measures were most stringent (47), supports this interpretation. The concentration of the mortality deficit in the younger age groups also argues against an important role of a reduced prevalence of other pathogens. For example, influenza leads to mortality in the older age groups, the lack thereof would therefore be expected to result in a mortality deficit in these age groups. In any case, we find no evidence for an overall detrimental effect of control measures on mortality. However, we cannot exclude harmful effects such as delays or avoidance of medical care (6, 7), increases in substance use and suicidal ideation (8, 10) or increases in interpersonal violence (11).

Our results are not readily applicable to other countries. Switzerland is a high-income country with a relatively old but healthy population. The stringency of control measures was relatively mild compared to other European countries (48). While any harmful indirect effects of control measures appear to have been more than compensated in Switzerland, further research is required to quantify indirect effects in other countries. Our approach can be used for this purpose, but it does not elucidate the pathways leading to an increase or a decrease in mortality. Further research using cause-specific mortality data is needed to answer this question. Studies over many years are required to gauge the long-term effects on mortality, if any, of the COVID-19 pandemic.

In conclusion, shortcomings in testing coverage caused considerable underestimation of COVID-19-related deaths in Switzerland, particularly in older populations. Although COVID-19 control measures may have negative effects (e.g., delays in medical care or mental health impairments), we note that after subtracting deaths directly caused by SARS-CoV-2 infections, there were fewer deaths in Switzerland during the pandemic than expected. This deficit cannot be attributed to a displacement of mortality, as it was observed mainly in the 40-69 age group. This suggests that any negative effects of control measures on mortality were more than offset by the positive effects. These results have important implications for the ongoing debate about the appropriateness of COVID-19 control measures.

## Data Availability

Data on population and all-cause mortality is freely available on the FSO website at https://www.pxweb.bfs.admin.ch/pxweb/en/ and https://www.bfs.admin.ch/bfs/en/home/statistics/population/births-deaths.html. Data on laboratory-confirmed deaths is freely available online (https://www.covid19.admin.ch/en/overview). Daily mean temperature data is freely available online (https://cds.climate.copernicus.eu/cdsapp#!/dataset/reanalysis-era5-single-levels?tab=overview).

https://www.bfs.admin.ch/bfs/en/home/statistics/population/births-deaths.html

https://www.covid19.admin.ch/en/overview

https://cds.climate.copernicus.eu/cdsapp#!/dataset/reanalysis-era5-single-levels?tab=overview

## Acknowledgments

This study would not have been possible without the extraordinary efforts of the data science team at the Federal Office of Public Health. We also thank Rolf Weitkunat (Federal Statistical Office) for his helpful comments. This study was funded by the SFOPH and the Swiss National Science Foundation (grant 189498). CLA acknowledge funding from the EU’s Horizon 2020 research and innovation programme (project EpiPose, 101003688). GK is supported by an MRC Skills Development Fellowship [MR/T025352/1].

## Author Contributions

JR, AH and GK conceived the study. JR and GK drafted the first version of the manuscript, did all statistical analyses, and take responsibility for the integrity of the data and the accuracy of the data analysis. ME revised the manuscript. All authors contributed to the interpretation of data and read and approved the final manuscript.

