## Supplementary material for "Direct and indirect effects of the COVID-19 pandemic on mortality in Switzerland: A population-based study": SI Appendix

### List of Figures

S1 Association between weekly laboratory-confirmed COVID-19-related deaths and absolute excess mortality by phase. The black line shows the slope of association corresponding to a 1 to 1 relation. The red lines show the association estimated with the model (corresponding to the $\beta_1$ coefficients, the full line represents the point estimate and the dashed lines the lower and upper bounds of the 95% credible interval). . . . . . . . . . . . . . . . . . . . . . . . . . . . . . . . . . . . . . . . 13

S2 Mean and 95% credible intervals of $\beta_1$ (panel A) and $\beta_2$ (panel B) across the different cantons. The canton abbreviations are given in Table S2. . . . . . . . . . . . . . . . . . . . . . . . . . . . 14

### S1 Supporting Information Text

#### S1.1 Population models

##### S1.1.1 Model specification

Let $P_{ijkl}$ be the population for the i-th sex (male-female), j-th age-group (<40, 40-59, 60-69, 70-79, $\geq$ 80), k-th year (2010-2019) and l-th canton. Let $X_{1i}$ be the sex and $X_{2k}$ be the year covariate. To predict populations for the years 2020-2023, had the COVID-19 pandemic not occurred, we considered 6 models of the following formulation:

$$P_{ijkl} \sim \text{Poisson}(\mu_{ijkl})$$

$$\log(\mu_{ijkl}) = \beta_0 + \beta_1 X_{1i} + \beta_2 X_{2k} + b_{jkl}$$

where $b_{jkl}$ is a random effect that has an age, temporal and spatial structure, see Table 1. All random effects were assigned an iid structure: $w_j, v_k, u_l, \xi_{jkl}, \beta_{2jkl}, \sim N(0, \sigma_g^2), g = 1, \ldots, 5$ and for $\sigma_g^2$ we considered penalised complexity priors [1]. In particular, we selected a prior so that $\Pr(\sigma_t > 10) = 0.1$, implying that it is unlikely to have population counts larger than exp(10) based solely on the selected structure of the random effects. All the models were fit using the Integrated Nested Laplace Approximation (INLA) for quick computation [2].

| Model | Abbreviation | $b_{jkl}$ |
|---|---|---|
| 1 | BASE | $w_j + v_k + u_l$ |
| 2 | OV | $w_j + v_k + u_l + \xi_{jkl}$ |
| 3 | OV_INT | $w_j + v_k + u_l + w_j \otimes v_k + w_j \otimes u_l + v_k \otimes u_l + \xi_{jkl}$ |
| 4 | VC | $w_j + v_k + u_l + \beta_{2jkl} X_{2k}$ |
| 5 | VC_OV | $w_j + v_k + u_l + \xi_{jkl} + \beta_{2jkl} X_{2k}$ |
| 6 | VC_OV_INT | $w_j + v_k + u_l + w_j \otimes v_k + w_j \otimes u_l + v_k \otimes u_l + \xi_{jkl} + \beta_{2jkl} X_{2k}$ |

Table 1. Specification of the random effects of the different population models. The notation $\otimes$ defines the Kronecker product for the interaction between two random variables.

##### S1.1.2 Cross validation

To mimic the scenario we want to reproduce, we selected to leave out the past 3 years (as we want to predict 3 years of population) all-together and examine the model prediction. To compare model prediction we calculated the coverage proportion, root mean square error (RMSE) and the mean bias. The coverage proportion is defined as the probability that the true population value lies in the 95% model based credible intervals, the MSE as $E[(\tilde{Y}_{ijkl} - Y_{ijkl})^2]$ and the mean bias as $E[\tilde{Y}_{ijkl} - Y_{ijkl}]$, where $\tilde{Y}_{ijkl}$ are the predictions and $Y_{ijkl}$ the true values of the population.

###### S1.1.3 Results

Table 2, shows the results of the cross validation and it is apparent that model 3, performs best, and this was the model selected for the subsequent analysis of excess deaths.

| Model | Coverage | RMSE | mean bias |
|---|---:|---:|---:|
| BASE | 0.08 | 6198 | 15 |
| OV | 0.96 | 7481 | 145 |
| OV_INT | 0.98 | 6009 | 9 |
| VC | 0.38 | 7415 | 175 |
| VC_OV | 0.55 | 6265 | 17 |
| VC_OV_INT | 0.61 | 7694 | 139 |

Table 2. Coverage, root mean square error (RMSE) and mean bias for the different models based on the leave out the past 3 years cross validation scheme.

#### S1.2 Expected mortality model

##### S1.2.1 Model specification

We used a Bayesian hierarchical model to predict deaths in 2020-2022, under the counterfactual scenario of absence of the COVID-19 pandemic. Let $Y_{jtkl}$ be the number of all-cause deaths, $P_{jtkl}$ be the population at risk (for the weeks during 2010-2019 this is fixed, whereas we used 200 samples to propagate the model based uncertainty from the population model for the weeks during 2020-2022) and $r_{jtkl}$ the risk in the $j$-th age-sex group (male-female and <40, 40-59, 60-69, 70-79, $\geq$ 80), $t$-th week ($t = 1, \dots, 595$, with 1 denoting the first week of 2011 and 595 the week that ends on the 3[rd] of April), the $k$-th year ($k = 1, \dots, 13$ with year 1 corresponding to 2011) and $l$-th canton ($s = 1, \dots, S$). We have to note that unlike the previous model, we did not group by sex in this analysis, as previous analysis showed small differences across the different sexes [3]. We assume a Poisson distribution for the number of deaths $Y_{jtkl}$ and specified the following model:

$$Y_{jtkl} \sim \text{Poisson}\big(r_{jtkl} P_{jtkl}\big)$$

$$\log\left(r_{jtkl}\right) = \beta_{0t} + \beta_1 X_{1t} + \beta_2 X_{2j} + \beta_3 X_{3k} + f(x_{jtkl}) + v_t + u_l,$$

where $\beta_{0t}$ is a week specific intercept given by $\beta_{0t} = \beta_0 + \epsilon_t$, with $\beta_0$ being the global intercept and $\epsilon_t \sim \text{Normal}(0, \sigma^2_\epsilon)$ an unstructured random effect representing the deviation of each week from the global intercept, with $\sigma^2_\epsilon$ denoting the variance of $\epsilon_t$. The term $\beta_1$ represents the effect of public holidays, $\beta_2$ the effect of age and $\beta_3$ a linear term to capture the long-term trend of the mortality. The effect of (mean weekly and cantonal) temperature on the all-cause mortality is captured in the flexible functions $f(\cdot)$. We define $f(\cdot)$ as a second-order random walk:

$$x_{tl} \mid x_{(t-1)l}, x_{(t-2)l}, \sigma^2_x \sim \text{Normal}\left(2x_{(t-1)l} + x_{(t-2)l}, \sigma^2_x\right), \tag{1}$$

with $\sigma^2_x$ denoting the variance and $x_{tkl}$ the temperature in the $t$-th week of the $k$-th spatial unit.

We accounted for seasonality using a non-linear weekly effect $v_t$ with a first order random walk (RW1) structure:

$$v_t \mid v_{t-1}, \sigma^2_v \sim \text{Normal}(v_{t-1}, \sigma^2_v),$$

where $\sigma^2_v$ is the variance of $v_t$.

The term $u_l$ is defined as a reparametrisation of the Besag-York-Mollié model given by the sum of an unstructured random effect, $\gamma_l \sim \text{Normal}(0, \sigma^2_\gamma)$, and a spatially structured effect $\delta_l$ [4, 5]. In particular $u_l$ is defined as:

$$u_l = \sigma^2_u \left(\sqrt{1-\theta}\gamma^\star_l + \sqrt{\theta}\delta^\star_l\right),$$

where $\gamma_l^\star$ and $\delta_l^\star$ are standardised version of $\gamma_l$ and $\delta_l$ to have variance equal to 1 [1]. The term $0 \leq \theta \leq 1$ is a mixing parameter which measures the proportion of the marginal variance explained by the structured effect and $\sigma_u^2$ the variance of the spatial field.

##### S1.2.2 Prior specification

We specified uninformative priors for the fixed effects $\beta_0$, $\beta_1$ and $\beta_2$. For the hyperparameters of the random effects we considered priors that tend to regularise inference while not providing too strong information [1]. For the standard deviation of the spatial field we defined a prior so that $\Pr(\sigma_u > 1) = 0.01$, implying that it is unlikely to have a spatial relative risk higher than exp(2) based solely on spatial variation. For $\theta$ we set $\Pr(\theta < 0.5) = 0.5$ reflecting our lack of knowledge about which spatial component, the unstructured or structured, should dominate the field $u$. For the rest of the standard deviations we defined the priors as $\Pr(\sigma_* > 1) = 0.01$.

##### S1.2.3 Cross validation

To examine the predictive ability of the above-mentioned model we defined a cross validation scheme, leaving the past 2 years out (we did not leave a third year out, as our predictions for 2022 are only for the first 3 months). We used metrics regarding the coverage proportion, bias, and correlation between predicted and true value of deaths. The results of the cross validation are shown on Figures 1-5 and Table 3.

Figure 1 shows the correlation between predicted and true value of deaths by age, sex and year (across the weeks and cantons). We see that the correlation is highest for the older groups and is not modified by year or sex. Figure 2 shows the bias by age, sex and year (across the weeks and cantons) and we see that it is always centred in 0. Figure 3 shows the coverage proportion by age, sex and year and we see that is pretty high for all groups, with the lowest (0.95) being for the older age groups. We also calculated the relative bias (truth-predicted / truth) by canton and and week and we can see that there the model is overall unbiased, Figure 4 and 5. Table 3 calculated the relative bias in the totals by age, sex in 2018, 2019 and 2018 and 2019 combined. We observed higher relative bias in females in general, but the overall relative bias for the totals is 0.

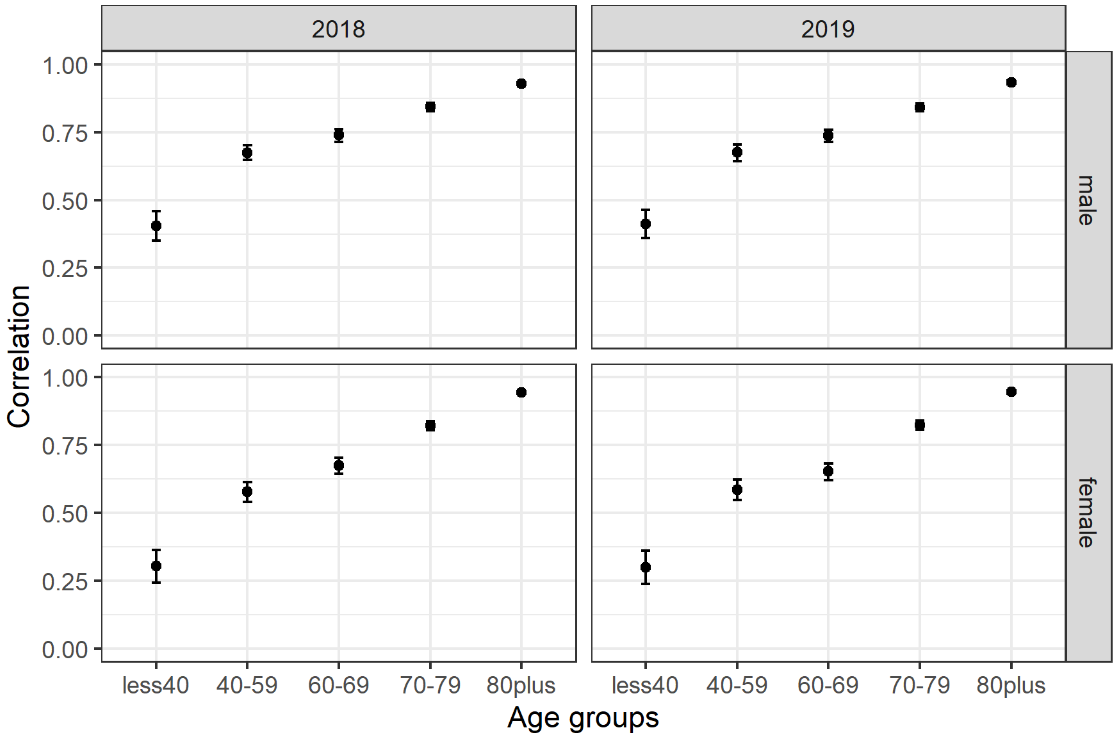

Figure 1. Correlation between the predicted and observed weekly and cantonal number of deaths by year, age and sex.

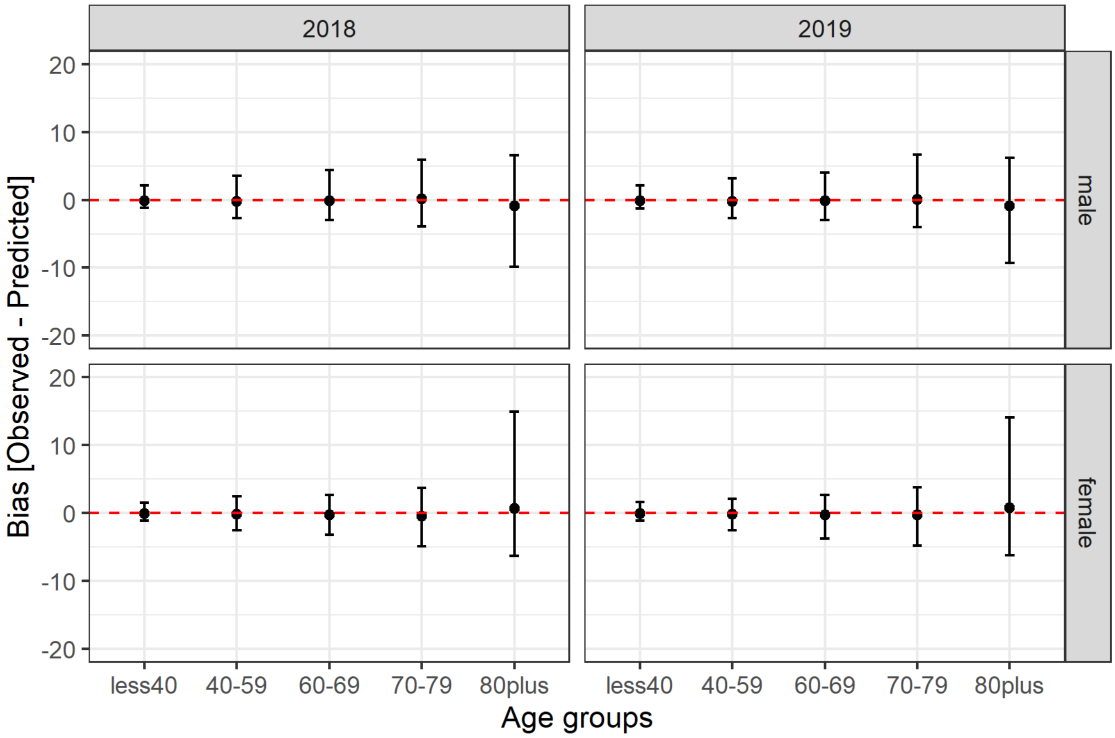

Figure 2. Bias between the predicted and observed weekly and cantonal number of deaths by year, age and sex.

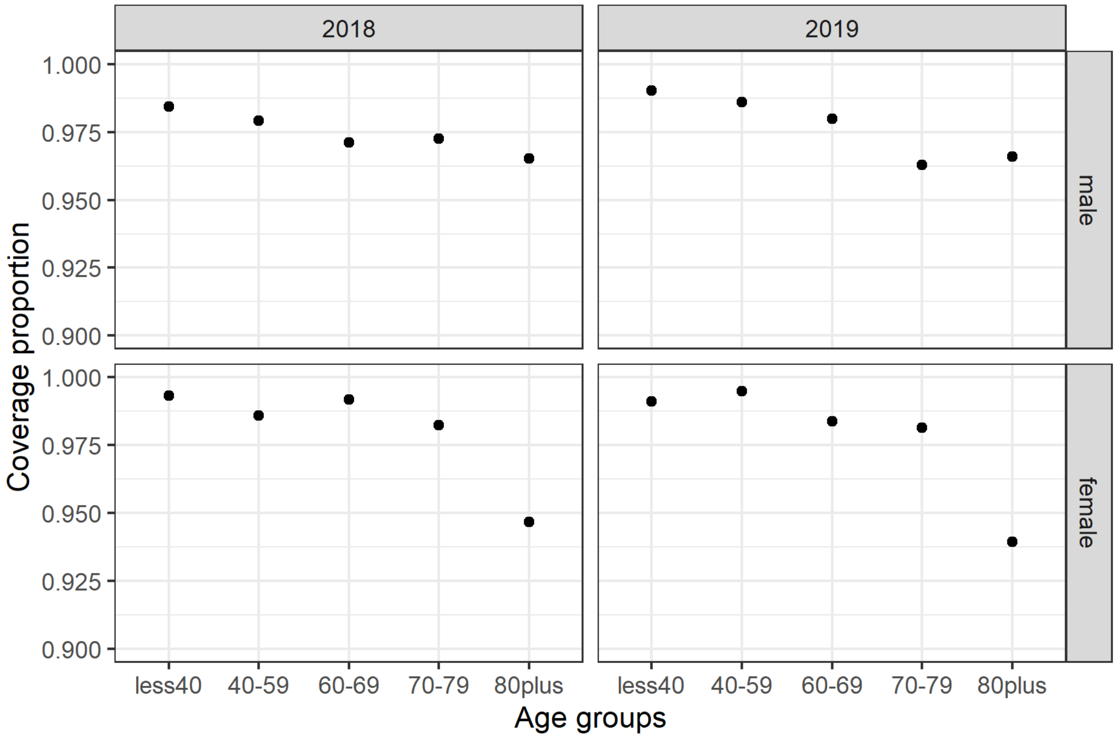

Figure 3. Coverage proportion by year, age and sex.

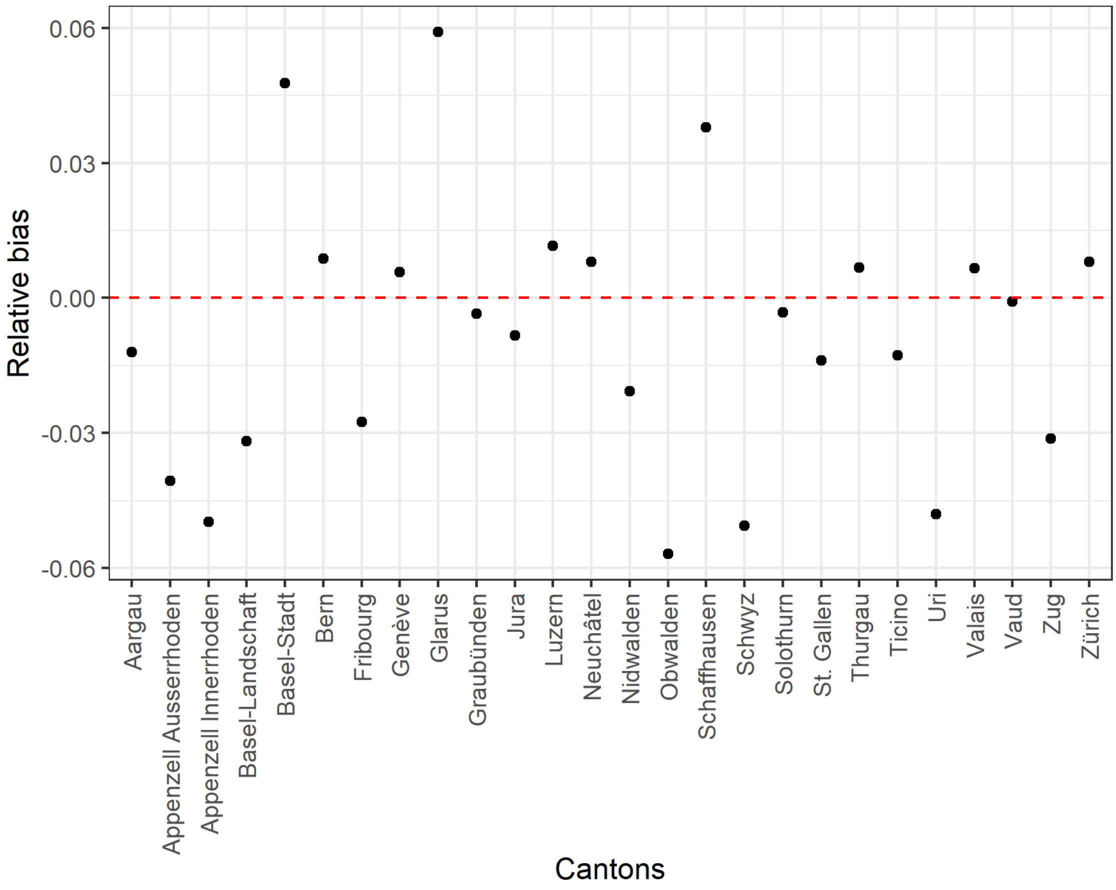

Figure 4. Relative bias by canton.

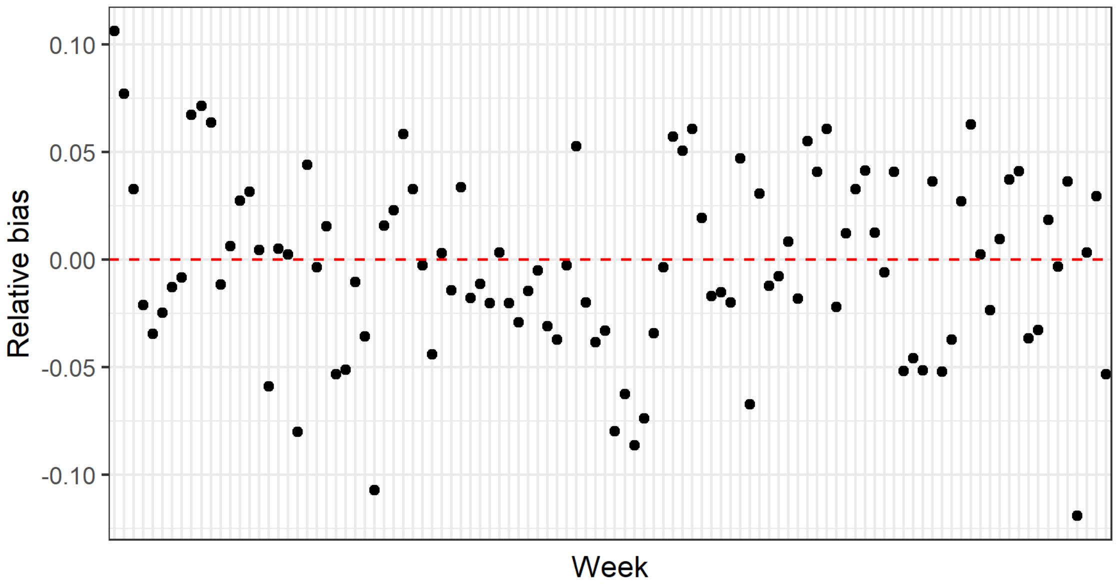

Figure 5. Relative bias by week.

| age | sex | 2018 | 2019 | 2018-2019 |
|---|---|---|---|---|
| less40 | male | 0.05 | 0.01 | 0.04 |
| 40-59 | male | 0.04 | 0.00 | 0.02 |
| 60-69 | male | 0.07 | 0.04 | 0.05 |
| 70-79 | male | 0.06 | 0.07 | 0.07 |
| 80plus | male | -0.09 | -0.10 | -0.10 |
| less40 | female | -0.13 | -0.22 | -0.18 |
| 40-59 | female | -0.11 | -0.19 | -0.15 |
| 60-69 | female | -0.16 | -0.19 | -0.17 |
| 70-79 | female | -0.11 | -0.08 | -0.10 |
| 80plus | female | 0.08 | 0.09 | 0.08 |
| less40 | Total | -0.02 | -0.07 | -0.04 |
| 40-59 | Total | -0.01 | -0.07 | -0.04 |
| 60-69 | Total | -0.02 | -0.05 | -0.03 |
| 70-79 | Total | -0.01 | 0.01 | -0.00 |
| 80plus | Total | 0.01 | 0.01 | 0.01 |
| Total | Total | 0.00 | -0.00 | -0.00 |

Table 3. Relative bias by age, sex and year.

#### S1.3 Decomposition model

We used a Bayesian hierarchical models to evaluate the association between all-cause deaths and laboratory-confirmed COVID-19-related deaths and expected all-cause mortality had the pandemic not occurred. Let $O_{tk}$ be number all-cause deaths, $L_{tk}$ the laboratory-confirmed COVID-19-related deaths and $E_{tkq}$ the expected all-cause mortality had the pandemic not occurred, on the $t$-th week for the $k$-th group for the $q$-th sample (the group can be either one of the following: Total, age group, phase and cantons). Then the model is specified as follows:

$$\begin{aligned}
O_{tk} &\sim \text{Poisson}(\lambda_{tk}) \\
\lambda_{tk} &= \sum_k \beta_{1k} L_{tk} + \sum_k \beta_{2k} E_{tkq} + u_{tk} \\
u_{tk} &\sim \text{Normal}(0, \sigma_u^2), \text{ with } \sum_{tk} u_{tk} = 0 \\
\beta_{11}, \dots \beta_{1k} &\sim \text{Normal}(\beta_1, \sigma_1^2) \\
\beta_{21}, \dots \beta_{21} &\sim \text{Normal}(\beta_2, \sigma_2^2) \\
\beta_1 &\sim \text{Normal}(0^+, 2) \\
\beta_2 &\sim \text{Normal}(0^+, 5) \\
\sigma_1^2 &\sim \text{Exponential}(r_1 = 1) \\
\sigma_2^2 &\sim \text{Exponential}(r_1 = 0.1) \\
\sigma_u^2 &\sim \text{Exponential}(r_1 = 0.001)
\end{aligned}$$

$\beta_{11}, \dots \beta_{1k}$ and $\beta_{21}, \dots \beta_{2k}$ are the effects of laboratory-confirmed COVID-19-related deaths and expected all-cause mortality had the pandemic not occurred for the k-th subgroup. We used a multi-level structure on the betas, providing some level of smoothing by introducing a global mean. The global mean for $\beta_{11}, \dots \beta_{1k}$ is $\beta_1$ and comes from a truncated normal with 0 mean and 2 variance, whereas for for $\beta_{21}, \dots \beta_{2k}$ is $\beta_2$ and comes from a truncated normal with 0 mean and 5 variance. $u_{tk}$ is an overdispersion term accounting for extra-Poisson variability and comes from a normal distribution with 0 mean and variance $\sigma_u^2$. Notice, that we impose sum-to-zero constraints for $u_{tk}$ making sure the model has no intercept. All variance hyperparameters were assigned vague exponential priors with varying rates ($r$) depending on the scale of each random variable.

We repeat the above procedure for $q = 1, \dots, 200$, pooled the samples and took a random sample of 1000 realisations from the pooled sample.

Table S1: Mean and 95% credible intervals of $\beta_1$ and $\beta_2$ for the Total group, the different phases and age groups considered.

| | $\beta_1$ | | | $\beta_2$ | | |
|---|---|---|---|---|---|---|
| | Mean | 2.5% | 97.5% | Mean | 2.5% | 97.5% |
| Total | 1.38 | 1.22 | 1.54 | 0.97 | 0.93 | 1.01 |
| Phase 1 | 1.46 | 1.03 | 1.87 | 0.95 | 0.89 | 1.00 |
| Phase 2 | 2.50 | 0.28 | 7.49 | 0.98 | 0.93 | 1.03 |
| Phase 3 | 1.46 | 1.28 | 1.67 | 0.93 | 0.85 | 1.00 |
| Phase 4 | 0.62 | 0.02 | 1.73 | 0.94 | 0.87 | 1.01 |
| Phase 5 | 2.61 | 0.76 | 5.07 | 0.99 | 0.92 | 1.07 |
| Phase 6 | 2.30 | 1.34 | 3.21 | 0.99 | 0.91 | 1.09 |
| Phase 7 | 1.70 | 0.55 | 2.90 | 0.95 | 0.86 | 1.04 |
| Age group: 0-39 | 1.34 | 0.19 | 3.27 | 0.94 | 0.82 | 1.05 |
| Age group: 0-59 | 1.01 | 0.13 | 1.76 | 0.88 | 0.81 | 0.96 |
| Age group: 60-69 | 1.11 | 0.52 | 1.56 | 0.91 | 0.84 | 0.99 |
| Age group: 70-79 | 1.20 | 0.99 | 1.41 | 1.01 | 0.95 | 1.07 |
| Age group: $\geq 80$ | 1.48 | 1.37 | 1.60 | 0.98 | 0.93 | 1.03 |

Table S2: Mean and 95% credible intervals of $\beta_1$ and $\beta_2$ across the different cantons.

| | | $\beta_1$ | | | $\beta_2$ | | |
|---|---|---|---|---|---|---|---|
| Canton | Abbreviation | Mean | 2.5% | 97.5% | Mean | 2.5% | 97.5% |
| Aargau | AG | 1.39 | 1.18 | 1.58 | 0.99 | 0.92 | 1.07 |
| Appenzell Innerrhoden | AI | 1.47 | 1.16 | 1.89 | 0.95 | 0.84 | 1.07 |
| Appenzell Ausserrhoden | AR | 1.42 | 1.11 | 1.74 | 0.93 | 0.83 | 1.03 |
| Bern | BE | 1.29 | 1.07 | 1.50 | 0.95 | 0.87 | 1.05 |
| Basel-Landschaft | BL | 1.46 | 1.19 | 1.77 | 1.01 | 0.94 | 1.10 |
| Basel-Stadt | BS | 1.49 | 1.24 | 1.78 | 0.90 | 0.80 | 0.99 |
| Fribourg | FR | 1.40 | 1.19 | 1.59 | 0.98 | 0.90 | 1.07 |
| Geneve | GE | 1.39 | 1.21 | 1.55 | 0.95 | 0.86 | 1.03 |
| Glarus | GL | 1.43 | 1.12 | 1.78 | 0.94 | 0.84 | 1.04 |
| Graubunden | GR | 1.39 | 1.10 | 1.70 | 0.94 | 0.86 | 1.02 |
| Jura | JU | 1.62 | 1.29 | 2.10 | 1.01 | 0.93 | 1.11 |
| Luzern | LU | 1.47 | 1.22 | 1.76 | 0.99 | 0.92 | 1.09 |
| Neuchatel | NE | 1.31 | 1.06 | 1.52 | 0.89 | 0.81 | 0.98 |
| Nidwalden | NW | 1.47 | 1.13 | 1.89 | 0.97 | 0.87 | 1.08 |
| Obwalden | OW | 1.45 | 1.17 | 1.79 | 0.96 | 0.85 | 1.06 |
| St. Gallen | SG | 1.53 | 1.34 | 1.72 | 1.01 | 0.94 | 1.10 |
| Schaffhausen | SH | 1.44 | 1.15 | 1.78 | 0.97 | 0.88 | 1.06 |
| Solothurn | SO | 1.48 | 1.24 | 1.74 | 0.96 | 0.88 | 1.05 |
| Schwyz | SZ | 1.51 | 1.26 | 1.81 | 1.01 | 0.92 | 1.10 |
| Thurgau | TG | 1.34 | 1.14 | 1.53 | 0.95 | 0.88 | 1.03 |
| Ticino | TI | 1.35 | 1.19 | 1.50 | 0.96 | 0.89 | 1.06 |
| Uri | UR | 1.41 | 1.08 | 1.76 | 0.92 | 0.81 | 1.01 |
| Vaud | VD | 1.48 | 1.31 | 1.66 | 0.95 | 0.86 | 1.03 |
| Valais | VS | 1.23 | 1.05 | 1.41 | 0.95 | 0.87 | 1.04 |
| Zug | ZG | 1.60 | 1.32 | 2.03 | 0.97 | 0.88 | 1.07 |
| Zurich | ZH | 1.38 | 1.15 | 1.60 | 0.97 | 0.89 | 1.05 |

Figure S1: Association between weekly laboratory-confirmed COVID-19-related deaths and absolute excess mortality by phase. The black line shows the slope of association corresponding to a 1 to 1 relation. The red lines show the association estimated with the model (corresponding to the $\beta_1$ coefficients, the full line represents the point estimate and the dashed lines the lower and upper bounds of the 95% credible interval).

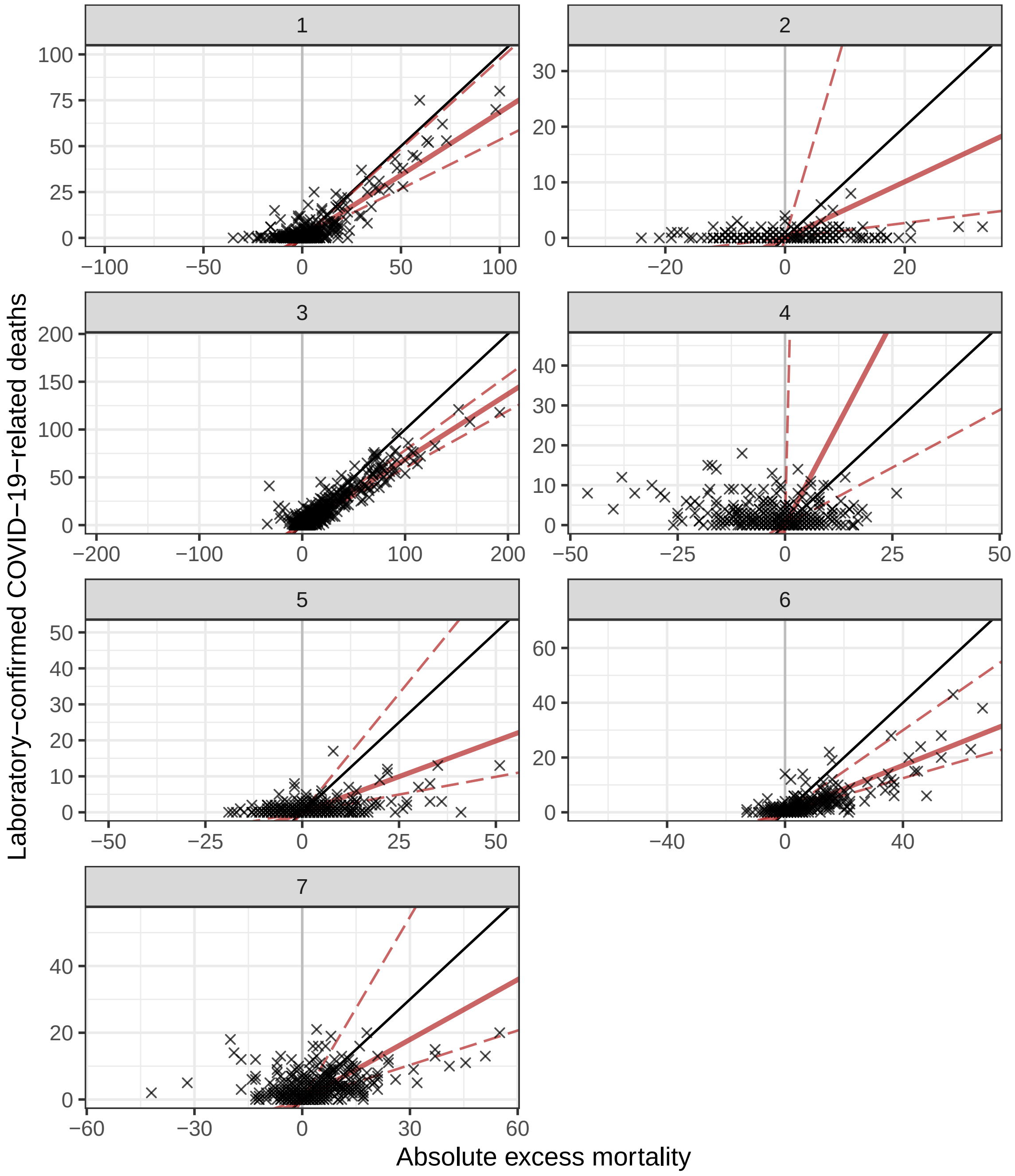

Figure S2: Mean and 95% credible intervals of $\beta_1$ (panel A) and $\beta_2$ (panel B) across the different cantons. The canton abbreviations are given in Table S2.

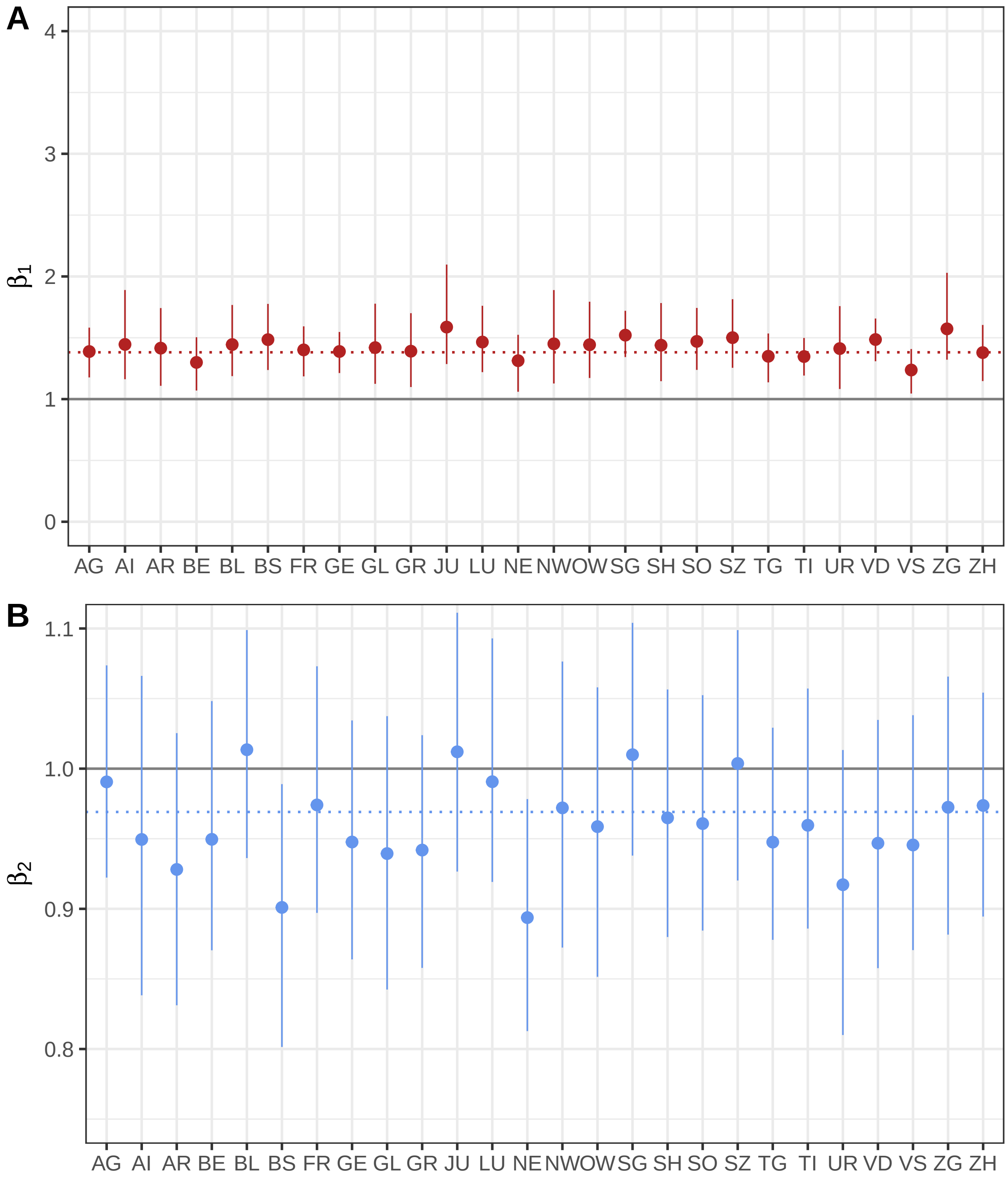
